# Performance of ChatGPT in pediatric audiology as rated by students and experts

**DOI:** 10.1101/2024.10.24.24316037

**Authors:** Anna Ratuszniak, Elzbieta Gos, Artur Lorens, Piotr H. Skarzynski, Henryk Skarzynski, W. Wiktor Jedrzejczak

## Abstract

**Background:** Despite the growing popularity of artificial intelligence (AI)-based systems such as ChatGPT, there is still little evidence of their effectiveness in audiology, particularly in pediatric audiology. The present study aimed to verify the performance of ChatGPT in this field, as assessed by both students and professionals, and to compare its Polish and English versions.

**Material and methods:** ChatGPT was presented with 20 questions, which were posed twice, first in Polish and then in English. A group of 20 students and 16 professionals in the field of audiology and otolaryngology rated the answers on a Likert scale from 1 to 5 in terms of correctness, relevance, completeness, and linguistic accuracy. Both groups were also asked to assess the usefulness of ChatGPT as a source of information for patients, in educational settings for students, and in professional work.

**Results:** Both students and professionals generally rated ChatGPT’s responses to be satisfactory. For most of the questions, ChatGPT’s responses were rated somewhat higher by the students than the professionals, although statistically significant differences were only evident for completeness and linguistic accuracy. Those who rated ChatGPT’s responses more highly were also rated higher it usefulness.

**Conclusions:** ChatGPT can possibly be used for quick information retrieval, especially by non-experts, but it lacks the depth and reliability required by professionals. The different ratings given by students and professionals, and its language dependency, indicate it works best as a supplementary tool, not as a replacement for verifiable sources, particularly in a healthcare setting.

## Introduction

With the proliferation of information and communication technologies, using the Internet to seek health information is common [1,2]. This approach is often preferred because of its availability and coverage, convenience, affordability, interactivity, and anonymity [3,4]. Seeking health information online enables a person to quickly learn more about their health problems, manage a health condition, decide about a health option, or change behavior [5]. Increasing numbers of patients are seeking health information online, making health information seeking behavior into an acronym (HISB) and a global trend. Chatbots based on large language models (LLMs) are becoming more commonly used for HISB [6].

One of the most advanced artificial intelligence (AI) language models is ChatGPT developed by OpenAI (San Francisco, California, USA) [7]. ChatGPT is an LLM that uses machine-learning techniques to generate human-like text based on a given prompt. Based on a large corpus of text, ChatGPT is able to capture the subtleties of human language, allowing it to generate appropriate and contextually relevant responses across a broad spectrum of topics [8]. Launched in November 2022, it quickly gained popularity and has become one of the fastest-growing web applications ever. According to the latest data, ChatGPT currently has approximately 180 million users [9].

While it appears that ChatGPT can provide significant support in many areas of science and education, there is potential for misuse, including the provision of biased content, limited credibility, creation of dishonest views and opinion, and others [1]. ChatGPT is currently being widely tested in various fields of knowledge, including science and medicine. The potential applications of ChatGPT in the medical field range from identifying potential research topics to assisting professionals in clinical diagnosis. It has been used in the fields of psychiatry, dermatology, ophthalmology, radiology, oncology, neurology, pharmacology, and others [10–16]. There are numerous studies in the field of otorhinolaryngology [6,17,18], but strangely there are only a handful in the related field of audiology and none focusing on pediatric issues [19–23]. This presents an unwelcome gap, since audiology deals with important issues surrounding the diagnosis, management, and treatment of hearing loss, as well as balance problems. An audiologist is responsible for fitting and dispensing hearing aids, providing hearing rehabilitation, and helping in the prevention of hearing loss. While hearing problems are usually not life-threatening, they have a not insignificant impact on society more generally. Unlike a major disease for which a patient will immediately seek help from a professional, it appears likely that a person with a minor hearing problem (or the parent of a child with such a problem) will first seek information from the Internet.

ChatGPT and other artificial intelligence tools in medicine can be used by different groups of users. One group is patients and their relatives, who are looking for information and support from the internet. Another group is students who may use this tool as an educational aid. Lastly, there are professionals who are looking for an advisory tool. Within otolaryngology, validation of information from the internet is important for all these groups, although their needs are qualitatively different. The real question is, how valid are AI-based information sources in this field? [6].

This study evaluates audiology information provided by ChatGPT in terms of the correctness, relevance, completeness, and linguistic accuracy of responses to a defined set of questions related to pediatric audiology. We also specifically wanted to know whether students and experts rated the responses similarly, and whether there were differences when questions were presented in English or Polish.

## Material and methods

The responses of ChatGPT version 3.5 to a series of questions were analyzed. The publicly available standard version was used, which has a setting by which the language of the dialog can be altered (English is the default but Polish is an option). A total of 20 questions related to pediatric audiology were prepared by the students and checked by the lecturer (Table 1). The questions focused on topics relating to: hearing aids (questions 1, 3, 12, 16, 20); diagnosis and audiological testing (5-7, 9-11, 13, 18); and diseases and treatment (2, 4, 8, 14, 15, 17, 19). One of the questions (number 15) was specific to Poland.

**Table 1.** English versions of the questions that were posed to ChatGPT (translated from Polish using DeepL).

| No | Question |
| --- | --- |
| 1 | What is the CROS system? |
| 2 | What is conductive hearing loss? |
| 3 | What frequencies are responsible for the reception of speech sounds in children? |
| 4 | What can be the causes of otitis in a child? |
| 5 | How to interpret the test of tonal audiometry, what is the norm of hearing? |
| 6 | What is the ABR test in a child? |
| 7 | What is newborn hearing screening? |
| 8 | What is the course of exudative otitis media in a child? |
| 9 | How can hearing be tested in a one-year-old child? |
| 10 | What is the verbal audiometry* test? |
| 11 | What types of tympanograms are distinguished in impedance audiometry? |
| 12 | For what purpose is an ear impression performed in children? |
| 13 | How to determine air conduction in the audiogram? |
| 14 | What should be done in the case of an abnormal result of hearing screening in a newborn? |
| 15 | What do the yellow and blue certificates in the child's health book mean in the context of hearing screening of children in Poland? |
| 16 | What are the contraindications to the use of air-conduction hearing aids? |
| 17 | What are the most effective methods of treatment of exudative otitis media in a child? |
| 18 | What is the result of a verbal audiometry* test and how to interpret it? |
| 19 | What are the results of otoscopy for otosclerosis? |
| 20 | What implantable hearing prostheses can be used in children? |

\* 'verbal audiometry' is a term made up by DeepL when translating from Polish to English; the usual term is 'speech audiometry'.

The questions were submitted to ChatGPT in Polish (using the Polish language setting) on January 12, 2024. The questions were then translated from Polish into English using DeepL [24] and presented again to ChatGPT using the English language setting on January 18, 2024 (a related study has tested ChatGPT responses posed on different days or weeks and has not found significant differences due to time [21]).

The answers given in English were again translated back into Polish with DeepL, a tool that has been given good reviews [25]. The reasoning here was that the evaluation would not depend on the English-language skill of the evaluators. In this way we could take the translation element out of ChatGPT and compare the performance of its Polish and English settings in answering the same question. This process also simulated the way a person who didn’t know English would use the app if they knew the English version provided more information.

The two versions of answers to the same question (one using the Polish language route and the other the English language route, with the second translated back into Polish), were given to participants for evaluation (see supplementary file for English version of all responses). The only information they were given was that these two responses were collected at two different points of time.

The quality of ChatGPT responses was evaluated by two groups of participants, students and experts, and here the framework established by Wang and Strong [26] was used. This framework has four quality (Q) categories: (1) Intrinsic Q consists of accuracy, objectivity, believability, and reputation; (2) Contextual Q consists of value-added, relevancy, timeliness, completeness, and an appropriate amount of data; (3) Representational Q consists of interpretability, ease of understanding, and representational consistency. Correctness (Intrinsic Q) was defined as the factual correctness of an answer and the absence of errors. Relevance (Intrinsic Q) rated how much an answer was related to the question. Completeness (Contextual Q) evaluated whether all important information was provided. Finally, linguistic accuracy (Representational Q) was assessed by whether the text sounded natural, whether there were any strange phrases or surprising words used, and whether the technical terms were properly used. A similar approach has been used by two other studies that have tested ChatGPT [27,28]. Correctness, relevance, completeness, and linguistic accuracy of the answers were rated using a 5-point Likert scale (1 = very unsatisfactory, 2 = unsatisfactory, 3 = neutral, 4 = satisfactory, and 5 = very satisfactory).

In addition, participants were asked 5 general questions about the usefulness of ChatGPT for patients as a source of information, for students in education, and for specialists in professional work (Table 2). Again, the participants were asked to give a score on a 5-point Likert scale (1 = very low, 2 = low, 3 = neither low nor high, 4 = high, 5 = very high).

**Table 2.** Questions asked of the participants relating to the usefulness of ChatGPT.

| Question no | Question |
| --- | --- |
| 1a | How do you rate the usefulness of the ChatGPT tool for patients as a source of information? |
| 1b | How do you rate the usefulness of the ChatGPT tool for students in education? |
| 1c | How do you assess the usefulness of the ChatGPT tool for specialists in consulting difficult, specialized cases? |
| 2 | How do you assess the possibility of using the ChatGPT tool in your professional work? |
| 3 | How do you assess the level of risk to the patient in using ChatGPT to obtain information? |

The ChatGPT responses were evaluated by a group of students (the same who compiled the list of questions) and a group of experts. The students (*n* = 20) were in the third year of a bachelor’s degree in audiophonology (similar to audiology and speech-language therapy in other countries). The experts (*n* = 16) consisted of professionals working in the field of audiology and otolaryngology, with many years of clinical and scientific experience, and comprised medical doctors (*n* = 2), hearing care professionals (*n* = 7), and scientists (*n* = 7). There were 10 who had 20 years or more of professional experience, 2 who had 13 years, 1 had 12 years, 2 had 10 years, and 1 had 5 years. Among them, 3 were professors, 5 had a PhD, and the others were medical doctors or had a master’s degree.

The study was approved by the bioethics committee of the Institute of Physiology and Pathology of Hearing (KB.IFPS: 2/2024).

### Statistical analysis

A mixed-design ANOVA (Analysis of Variance) with Bonferroni adjustment was employed to evaluate the differences in ratings of ChatGPT responses between the students and the experts (the between-subject factor) and between the Polish and English versions (the within-subject factor). Analyses were conducted both on the average ratings and for each of the 20 questions separately. A Mann–Whitney U-test was used to compare the usefulness of ChatGPT as perceived by the students and the experts. For some analyses, Pearson correlations were also calculated. A p-value below 0.05 was considered statistically significant. The analysis was conducted using IBM SPSS Statistics (v. 24).

## Results

### General overview of ChatGPT’s response ratings

The average ratings of ChatGPT’s responses in four dimensions (correctness, relevance, completeness, and linguistic accuracy), as assessed by both the students and the experts and for the Polish and English versions, are presented in Table 3, together with the results of the ANOVA analysis.

**Table 3.** Average ratings by students and experts of the Polish and English versions of all ChatGPT responses.

|  |  | Polish version |  | English version |  | Group effect | Language version effect | Interaction effect |
| --- | --- | --- | --- | --- | --- | --- | --- | --- |
|  |  | *M* | *SD* | *M* | *SD* | *F*; *p* | *F*; *p* | *F*; *p* |
| Correctness | Student | 4.02 | 0.70 | 3.98 | 0.45 | 3.81;<br>*p* = 0.059 | 0.54;<br>*p* = 0.469 | 0.01;<br>*p* = 0.914 |
|  | Expert | 3.72 | 0.41 | 3.66 | 0.43 |  |  |  |
| Relevance | Student | 4.17 | 0.64 | 4.11 | 0.41 | 0.21;<br>*p* = 0.648 | 3.62;<br>*p* = 0.066 | 0.69;<br>*p* = 0.411 |
|  | Expert | 4.03 | 0.59 | 4.00 | 0.56 |  |  |  |
| Completeness | Student | 3.84 | 0.73 | 3.67 | 0.48 | 5.00;<br>*p* = 0.032* | 1.23;<br>*p* = 0.274 | 0.02;<br>*p* = 0.887 |
|  | Expert | 3.45 | 0.45 | 3.92 | 0.55 |  |  |  |
| Linguistic accuracy | Student | 3.93 | 0.83 | 4.16 | 0.65 | 5.39;<br>*p* = 0.044* | 19.07;<br>*p* < 0.001** | 0.53;<br>*p* = 0.470 |
|  | Expert | 3.45 | 0.45 | 3.78 | 0.51 |  |  |  |

\* - *p* < 0.05, \*\* - *p* < 0.01

The general pattern was that ChatGPT’s responses received the highest ratings for Relevance (at least 4 points on average), whereas the lowest ratings were for Completeness. The experts were generally more critical than the students, while both groups gave about equal ratings for the two language versions. The ANOVA revealed some statistically significant differences in the ratings given by students and experts, as well as between the Polish and English versions (bold numbers in Table 3).

The most pronounced effects were found for the linguistic accuracy of ChatGPT’s responses. The group effect was statistically significant, indicating that experts (M = 3.61; SD = 0.61) rated ChatGPT’s responses significantly lower than students (M = 4.04; SD = 0.61), regardless of the language version. This effect was moderate, *η*^2^ = 0.11. The language version effect was also statistically significant, indicating that ratings for the English version (M = 3.99; SD = 0.61) were significantly higher than for the Polish version (M = 3.71; SD = 0.72), regardless of whether the rater was a student or an expert. This effect was large, *η*^2^ = 0.36.

A statistically significant difference between students and experts was also found for the Completeness of ChatGPT’s responses. Again, experts (M = 3.48; SD = 0.54) rated ChatGPT’s responses significantly lower than students (M = 3.88; SD = 0.53), regardless of the language version. This effect was moderate, *η*^2^ = 0.11. Similar results were found for Correctness, although the effect did not reach statistical significance (p = 0.059). Experts (M = 3.69; SD = 0.47) rated ChatGPT’s responses generally as less correct than students (M = 4.00; SD = 0.48).

For Relevance, there was a tendency for ratings to be slightly higher for the Polish version (M = 4.11; SD = 0.61) than for the English version (M = 3.99; SD = 0.47), regardless of whether the rater was a student or an expert; however, this effect did not reach statistical significance (*p* = 0.066).

### Question-specific analysis of ChatGPT’s response ratings

ANOVA was also applied to the ratings provided by the students and the experts to both language versions of ChatGPT’s responses to each of the 20 questions. The colours in Table 4 indicate whether the ratings were higher for students (red) or experts (green). Saturated colors indicate statistically significant differences, while lighter shades represent non-significant differences.

**Table 4.**
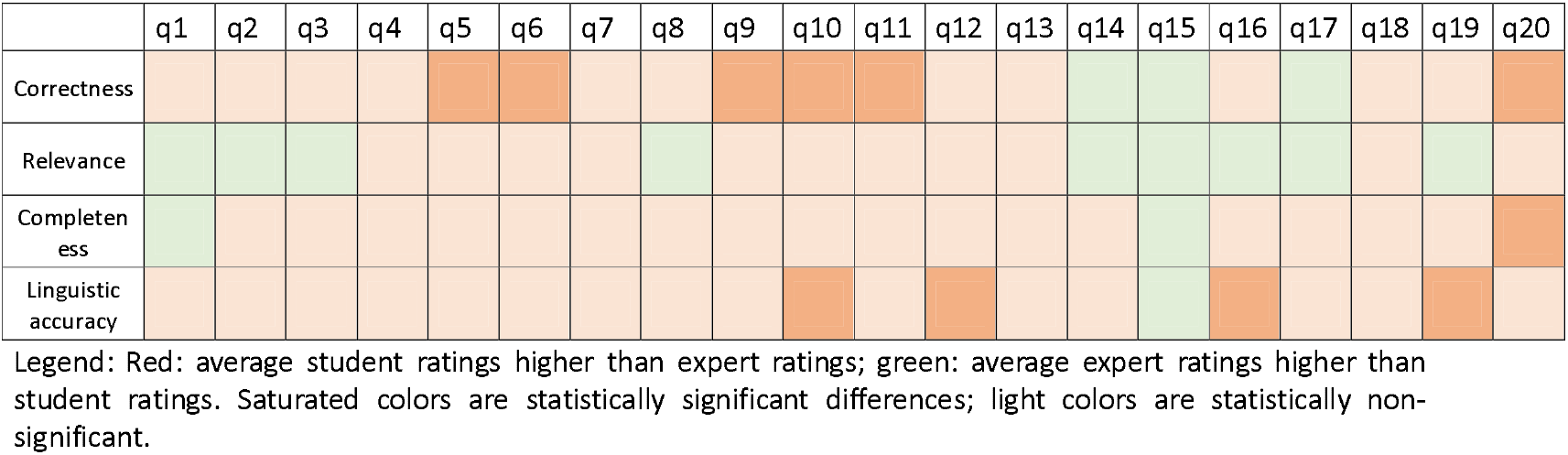
Comparison of students’ and experts’ ratings for ChatGPT’s responses across 20 questions (q1–q20).

The trend was that students generally rated ChatGPT’s responses higher than the experts across all questions (except question 15) and all evaluation dimensions. Experts tended to be more critical regarding quality of responses, particularly concerning linguistic accuracy and completeness.

Table 5 shows whether the ratings were higher for ChatGPT’s responses in the English version (in blue) or in the Polish version (in yellow). Once more, the use of saturated colours indicates that the observed differences are statistically significant, whereas lighter shades represent non-significant differences.

**Table 5.**
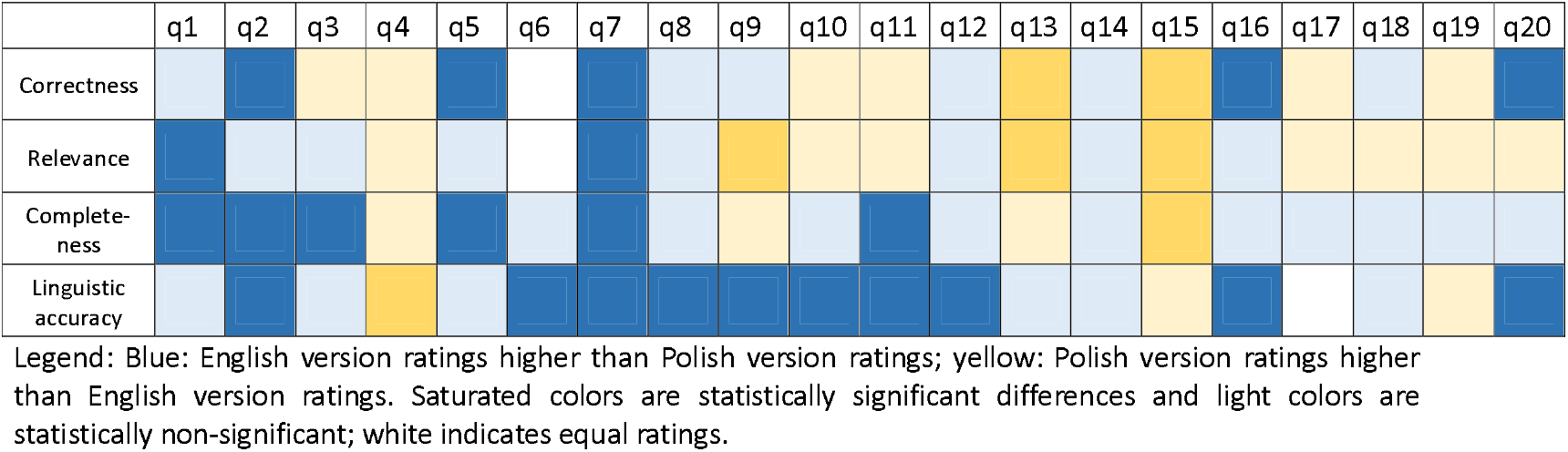
Comparison of English and Polish versions in terms of ratings of ChatGPT’s responses to 20 questions.

The overall observation is that the English version was rated higher than the Polish version. This is particularly evident in the context of linguistic accuracy and completeness. Again, an exception was the rating of responses to question 15, in which the Polish version was rated higher than the English version consistently across all dimensions.

One question, question 15, related particularly to Polish circumstances. It asked, What do the yellow and blue certificates signify in the context of hearing screening for children in Poland? For the Polish version, ChatGPT provided an explanation, whereas in the English version it responded: “I do not have specific information about the color-coding system used in child health books in Poland for hearing screening certificates” and followed with some advice about where such information could be found (see supplementary files for complete ChatGPT responses).

We examined how the ChatGPT responses to this question, both in the Polish and English versions, were rated by the students and the experts. The results are shown in Table 6.

**Table 6.**
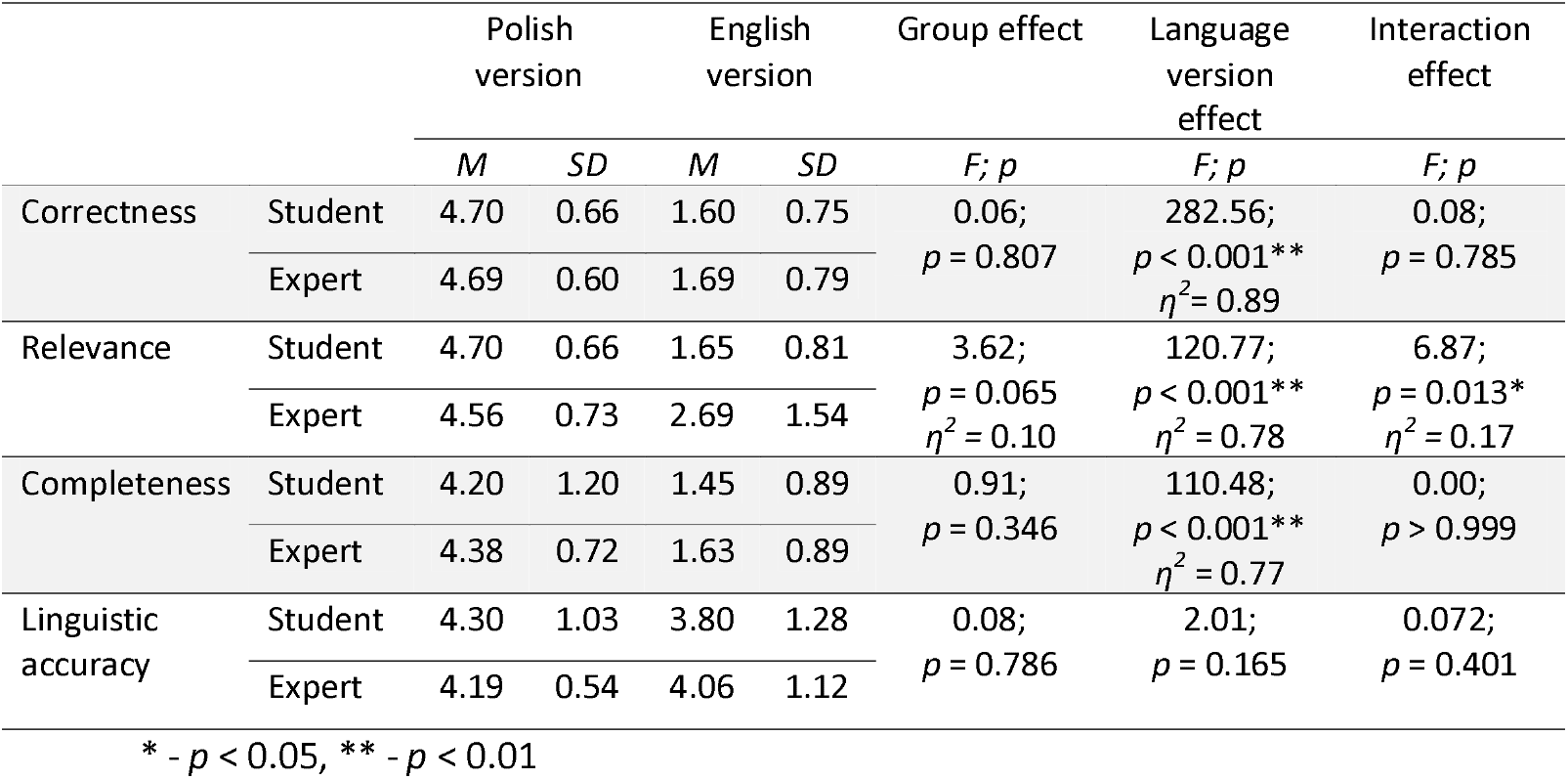
Ratings, by students and by experts, to ChatGPT’s responses to the Polish and English versions of question 15: *What do the yellow and blue certificates mean in the context of hearing screening for children in Poland?*

There were large differences between the ratings of the Polish and English versions of question 15 in terms of correctness, relevance, and completeness. However, the differences between the ratings by students and by experts of question 15 were not statistically significant (although in terms of Relevance, the difference almost reached significance, *p* = 0.065). The interaction effects (group x language version) were significant only in terms of Relevance.

For Correctness, there was a significant language effect in that the Polish version (M = 4.69; SD = 0.62) was generally rated higher than the English version (M = 1.64; SD = 0.76).

Similarly, a language effect was evident in the Relevance domain, where the Polish version (M = 4.64; SD = 0.68) was rated higher than the English version (M = 2.11; SD = 1.28). A significant interaction effect was that the experts rated ChatGPT’s responses significantly higher in the Polish version than in the English version (*p* < 0.001). The same was true for the students. A difference between the experts and the students was found only in the English version, which was rated significantly higher by the experts than the students (*p* = 0.014), although the ratings for the Polish version were similar across both groups.

For Completeness, a significant language effect was that ratings for the Polish version (M = 4.28; SD = 1.00) were significantly higher than for the English version (M = 1.53; SD = 0.88), irrespective of whether the rater was a student or an expert.

To sum up, ChatGPT’s responses to the specifically Polish question were, in the English version, consistently rated as less correct, less relevant, and less in-depth compared to the Polish version. Only the formal aspect of ChatGPT’s responses to this question, namely linguistic accuracy, was rated similarly across groups and language versions.

### Assessment of the usefulness of ChatGPT

Both students and experts evaluated the usefulness of ChatGPT in education, specialized fields, and patient care (Table 2). They also assessed its potential application in professional work and the associated risks of using ChatGPT for obtaining medical information.

Their assessments were compared using a Mann–Whitney U-test and are presented in Table 7.

**Table 7.** Ratings by students and experts of ChatGPT’s usefulness from Table 2.

| Question no | All participants | | Students | | Experts | | $U$; $p$ |
| --- | --- | --- | --- | --- | --- | --- | --- |
| | $M$ | $SD$ | $M$ | $SD$ | $M$ | $SD$ | |
| 1a | 3.78 | 0.72 | 3.90 | 0.72 | 3.63 | 0.72 | 120.0; $p = 0.160$ |
| 1b | 3.25 | 0.97 | 3.55 | 0.89 | 2.88 | 0.96 | 96.0; $p = 0.031^*$ |
| 1c | 2.06 | 0.83 | 2.20 | 0.89 | 1.87 | 0.72 | 127.0; $p = 0.264$ |
| 2 | 2.86 | 0.80 | 3.05 | 0.83 | 2.63 | 0.72 | 117.0; $p = 0.143$ |
| 3 | 2.94 | 0.75 | 3.00 | 0.86 | 2.88 | 0.62 | 151.0; $p = 0.753$ |

\* - $p < 0.05$

The ratings (on a scale of 1 to 5) shown in Table 7, and reflect differing levels of perceived usefulness of ChatGPT. Overall, the students rated ChatGPT more favorably than the experts. Notably, the students gave significantly higher ratings for its usefulness in education (question 1b) compared to the experts. While most other assessments were similar between the two groups, both rated ChatGPT relatively lower for consulting specialized cases (question 1c) and higher as a source of information for patients (question 1a). Ratings for potential professional use (question 2) and the perceived risk to patients (question 3) were moderate across both groups.

Finally, Table 8 combines the scores evaluating the correctness, relevance, completeness, and linguistic accuracy together with assessment of the usefulness of ChatGPT. The table presents correlations of average scores from evaluation of questions in both languages (since there were no language-specific correlations). The correlations in Table 8 are for the whole group of 36 evaluators (20 students and 16 experts). There were several significant positive correlations for correctness, completeness, and linguistic accuracy; however, relevance was not significantly correlated with any usefulness score.

**Table 8.**
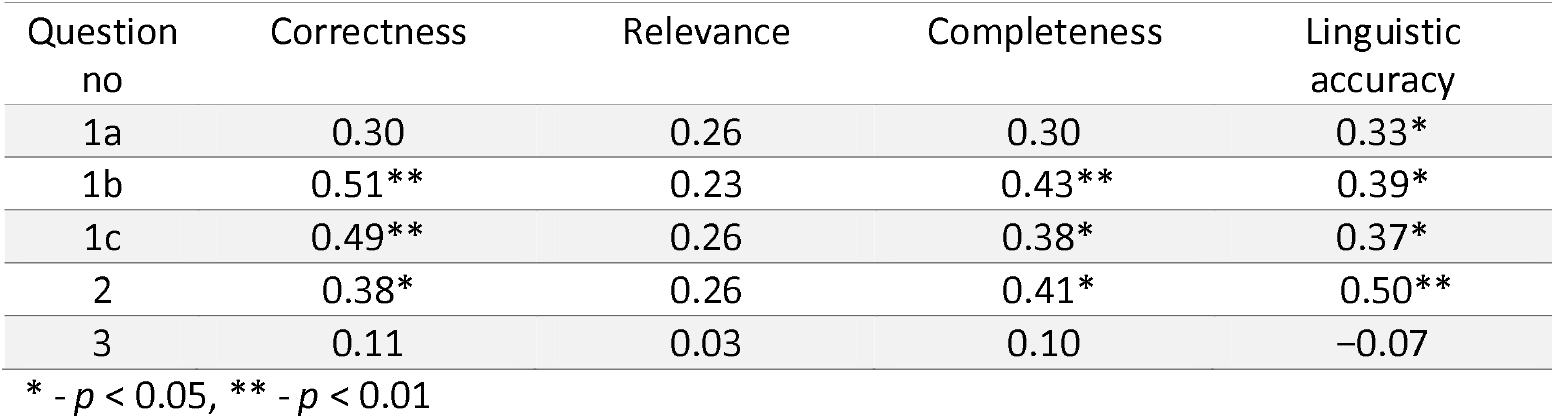
Correlation between the usefulness of ChatGPT and average ratings given to ChatGPT responses. The correlations are for the whole group of 36 evaluators (20 students and 16 experts). * - p < 0.05, ** - p < 0.01

## Discussion

This evaluation of the quality of ChatGPT responses to a set of questions related to topics in audiology brought out some interesting perspectives. Generally, the responses given by both students and experts were positive, although it should be underlined that they were mostly at the level of ‘satisfactory’, not ‘very satisfactory’. This close to ‘satisfactory’ rating applied to all categories of quality (intrinsic, contextual, and representational). Furthermore, the experts were generally more critical and gave lower ratings for completeness. Even though the responses to questions posed to ChatGPT in English were generally rated slightly higher by both students and experts, the response to question 15 (specific to Polish circumstances) was rated higher when questions were posed in Polish.

The long experience and large knowledge-base of a group of experts are expected to be important factors in how the answers provided by ChatGPT are evaluated. Experts are likely to be more critical than students. This study largely confirms this view, in that in almost all cases the experts’ evaluations were lower than those of the students, although the differences were not large enough to be statistically significant in every category analyzed. One might assume that if the answers given were exemplary (e.g. as from an encyclopedia), then the expert and student evaluations would be very high and be similar for both groups. If, however, there were differences, one might assume that they were due to greater knowledge and experience, which allows for quicker and easier recognition of shortcomings and ambiguities. The less someone knows about a particular field, the more they will be impressed by a well-formulated ChatGPT answer. We think this could be particularly harmful to patients who could be easily misled by incomplete or imprecise information.

The level of knowledge of ChatGPT in audiology and laryngology was rated as quite high, although the mean scores of the responses were closer to 4 (satisfactory) than 5 (very satisfactory), even in the student group. Although the differences in responses between the students and the experts were mostly not statistically significant, they show that students always gave higher scores than the experts. In general, the answers were pertinent and correct, but not without errors, so some caution should be exercised in their use.

A statistically significant difference in responses between the groups was found for the completeness category, which indicates whether all important information was given and whether the answer was too vague or superficial.

When one looks at the relevance category, it is here that differences due to knowledge and experience are most likely to be evident. Given that ChatGPT’s responses were of reasonably good quality, albeit not error-free, ChatGPT can be considered as a useful tool for people who wish to obtain quick information in a casual situation (e.g. overhearing something and wanting to check it). However, the results here show we should have limited trust in AI, and that patients should check information with verified sources. It is noteworthy that ChatGPT has a disclaimer on the bottom of the screen warning that errors are possible and information should be verified.

This study has shown that linguistic correctness is important, with experts being more critical than students in this area. This is probably because they can more readily navigate through the professional terminology, based on their own daily experience, whereas students only have the knowledge acquired from their teachers. As a result, experts are more likely to spot any inaccuracies or imprecise wording. Experts were easily able to identify common errors in ChatGPT responses such as lack of precision, colloquial word clusters, use of technical terms but in the wrong context, gibberish, generalizations, and truisms.

Comparison of the two language versions (Polish and English) showed that the quality of answers was better with the English setting than with the Polish setting. We were curious to find out whether the Polish and English versions of answers would be the same. Since we were not native speakers and not competent to qualitatively assess English texts, we used the DeepL tool (also based on AI) for translation into Polish. In most cases (the exception being question 15), the translated answers as given by DeepL were rated significantly better than the responses ChatGPT gave to the questions originally provided in Polish. English is the most common language by which ChatGPT has been trained, and studies have shown that it performs better in English than in other languages [11]. Nevertheless, this result is quite surprising in that despite two steps of translation (one Polish to English to frame the questions, and a second English to Polish to consistently frame responses) the responses were still rated as better than staying within a common Polish framework. At this stage we cannot give explain whether this is due to the better performance of ChatGPT in English or the translational skills of DeepL or both. However, this aspect is worth testing further, since translation tools are freely available on the internet.

At this point, question 15 is worth mentioning as it reflects a Polish-specific question (What do the yellow and blue certificates mean in the context of hearing screening for children in Poland?). In this case, ChatGPT’s response in Polish was rated higher than in English. This shows how care needs to be taken in asking questions about country-specific questions (and other cultural, religious, and linguistic circumstances) and how the language version used can affect the answer supplied. While the English version generally performed slightly better in our case, for certain local topics the native language version can clearly be better.

Both experts and students rated ChatGPT as useful for patients as a source of information (on average coming close to a ‘high’ rating). However, it rated lower for students in training (‘neither low nor high’), and was rated worst by professionals for consulting on difficult cases (‘low’). Two recent papers also note that AI might be more appropriate for gaining patient information than for training health professionals [29,30]. For all our categories, students rated the usefulness of ChatGPT more highly, although statistically significant differences between students and experts only showed up for the usefulness of ChatGPT as a teaching aid. Here the students’ ratings were statistically significantly higher than those of the experts. This may be because experts have a greater level of caution and skepticism towards new technologies, in contrast to the familiarity of searching online for information in the student group [30]. Nevertheless, caution is needed, as ChatGPT has a tendency to provide incorrect or non-existent scientific references, a risk that has been explicitly set out in the fields of education, research, and healthcare [30–32]. In our setting, both experts and students gave a neutral assessment of the usefulness of ChatGPT for their own professional work. Nevertheless, the participants who rated ChatGPT’s responses more highly were also more eager to use it professionally or as an education tool (Table 8).

This study sheds light on what categories of quality of health information are most important for people seeking information on the internet. As shown in Table 8, only ratings in the category of linguistic accuracy (Representational Q) correlated with ratings of patient usefulness. This suggests that both experts and students consider linguistic accuracy as highly important for patients, helping them interpret and understand health information. On the other hand, the strong correlations found in all categories of quality show that no category seems to be more important than another for students in education and for specialists in consulting difficult, specialized cases.

One important limitation of ChatGPT, and shared by all LLMs, is the phenomenon of hallucination – a seemingly realistic responses by an AI system which turn out to be incorrect [33]. This creates a potential risk to the patient in that they may be misled by false information. However, both students and experts saw little overall risk to patients from using ChatGPT in that way: there was no correlation between the assessment of risk to patients and any category of health information quality. None of the participants could point to any particularly harmful information in ChatGPT’s responses.

Finally, the major drawback of ChatGPT is that, even if directly asked, it does not provide sources of information or references to scientific papers, a crucial failing when involving scientific or medical knowledge [19]. This makes it impossible to verify any statement provided by ChatGPT, and, unless this drawback is overcome, it will remain the biggest limitation of this technology.

## Conclusions

While ChatGPT is a promising tool for initial information retrieval, particularly for non-experts, it falls short in providing the completeness and reliability required by professionals. While students found ChatGPT useful as an educational tool, experts were more skeptical, particularly since the program never provides verifiable references and sometimes provides misleading information. The differences in perception between students and experts, as well as between language versions, suggest that while AI can serve as a useful supplementary tool, it cannot replace traditional, verified sources, particularly in critical fields like healthcare.

## Data Availability

All data produced in the present study are available upon reasonable request to the authors

## Acknowledgment

The authors would like to thank the students and experts who participated in this study. The authors thank also Andrew Bell for comments on an earlier version of this manuscript.

