## Supplementary file for "Performance of ChatGPT in pediatric audiology as rated by students and experts"

SUPPLEMENTARY MATERIALS

### Questions to ChatGPT in English language version

1. What is the CROS system?
2. What is conductive hearing loss?
3. What frequencies are responsible for the reception of speech sounds in children?
4. What can be the causes of otitis in a child?
5. How to interpret the test of tonal audiometry, what is the norm of hearing?
6. What is the ABR test in a child?
7. What is newborn hearing screening?
8. What is the course of exudative otitis media in a child?
9. How can hearing be tested in a one-year-old child?
10. What is the verbal audiometry test?
11. What types of tympanograms are distinguished in impedance audiometry?
12. For what purpose is an ear impression performed in children?
13. How to determine air conduction in the audiogram?
14. What should be done in the case of an abnormal result of hearing screening in a newborn?
15. What do the yellow and blue certificates in the child's health book mean in the context of hearing screening of children in Poland?
16. What are the contraindications to the use of air-conduction hearing aids?
17. What are the most effective methods of treatment of exudative otitis media in a child?
18. What is the result of a verbal audiometry test and how to interpret it?
19. What are the results of otoscopy for otosclerosis?
20. What implantable hearing prostheses can be used in children?

### Questions with ChatGPT answers in English language version

1. **What is the CROS system?**

The CROS Hearing Aid System is a solution designed to help individuals with unilateral hearing loss. Unilateral hearing loss means that a person has significant hearing loss in one ear while the other ear has normal or near-normal hearing. The CROS system is particularly useful for those who have no usable hearing in one ear.

"CROS" stands for Contralateral Routing of Signals. Here's how the CROS Hearing Aid System typically works:

**Microphone on the Deaf Side:** A microphone is placed on the side of the head with hearing loss. This microphone picks up sounds from the deaf or impaired ear.

**Transmission to the Better Ear:** The captured sound is wirelessly transmitted to a hearing aid or receiver located on the better-hearing side. This allows the individual to hear sounds from both sides.

**Improved Awareness:** By transmitting sounds from the impaired side to the better-hearing ear, the CROS system helps individuals with unilateral hearing loss to be more aware of their surrounding environment and to better understand the directionality of sounds.

This technology is different from traditional hearing aids, which are designed to amplify sounds for individuals with hearing loss in both ears. The CROS Hearing Aid System is specifically tailored for those with one-sided hearing loss, providing a more comprehensive listening experience.

2. **What is conductive hearing loss?**

Conductive hearing loss is a type of hearing impairment that occurs when there is a problem with the transmission of sound waves from the outer or middle ear to the inner ear. This type of hearing loss can result from various issues that affect the ear's ability to conduct sound effectively.

Common causes of conductive hearing loss include:

Ear Infections: Infections in the ear canal or middle ear can cause inflammation and fluid buildup, affecting the movement of the eardrum and the small bones (ossicles) in the middle ear.

Blockage in the Ear Canal: Earwax (cerumen) buildup or the presence of a foreign object in the ear can block sound waves from reaching the eardrum.

Perforated Eardrum: A tear or hole in the eardrum can disrupt the normal transmission of sound.

Otosclerosis: Abnormal bone growth in the middle ear can interfere with the movement of the stapes bone, one of the three ossicles, leading to conductive hearing loss.

Malformation of the Ear: Some individuals may be born with structural abnormalities in the outer or middle ear that result in conductive hearing loss.

Conductive hearing loss is generally treatable, and in many cases, medical intervention or surgical procedures can help restore or improve hearing. Treatment options may include the removal of earwax, antibiotic treatment for infections, surgical repair of a perforated eardrum, or procedures to correct abnormalities in the middle ear.

It's important to note that conductive hearing loss is distinct from sensorineural hearing loss, which involves damage to the inner ear or auditory nerve and is often irreversible. In some cases, individuals may experience a combination of conductive and sensorineural hearing loss, known as mixed hearing loss. If you suspect hearing loss, it's advisable to consult with an audiologist or healthcare professional for a comprehensive hearing evaluation.

3. **What frequencies are responsible for the reception of speech sounds in children?**

Speech sounds cover a range of frequencies, and different frequencies contribute to the perception of various speech sounds. In general, the frequencies associated with speech sounds in children (as well as in adults) fall within the range of human hearing, which is approximately 20 Hz to 20,000 Hz. However, the key frequencies for speech understanding are typically found in the mid-frequency range.

The fundamental frequency (or pitch) of human speech, which is critical for conveying the intonation and emotional aspects of speech, usually falls between 125 Hz and 250 Hz. Vowels in speech are characterized by their formant frequencies, with the first formant typically ranging from around 300 Hz to 700 Hz and the second formant ranging from around 800 Hz to 1200 Hz.

Consonants, on the other hand, span a wider range of frequencies. Some of the higher-frequency components of speech associated with consonant sounds can extend beyond 2,000 Hz.

It's important to note that speech perception involves the integration of information across various frequencies, and the specific frequencies that contribute to the understanding of speech sounds can vary depending on the linguistic and acoustic characteristics of the language being spoken.

In the context of assessing and managing hearing loss in children, audiologists often focus on the frequency range associated with speech sounds to ensure that amplification devices (such as hearing aids) provide sufficient amplification in these critical frequency regions for effective speech comprehension and language development.

4. **What can be the causes of otitis in a child?**

Otitis, which refers to inflammation of the ear, can manifest in different forms. The two main types of otitis in children are otitis media and otitis externa, each with its own set of causes:

Otitis Media:

Bacterial or Viral Infection: Otitis media is often associated with infections caused by bacteria or viruses. Respiratory infections, such as the common cold, can lead to the accumulation of fluid in the middle ear, creating an environment conducive to bacterial or viral growth.

Eustachian Tube Dysfunction: The Eustachian tubes, which connect the middle ear to the back of the throat, help regulate air pressure in the ear. Dysfunction of these tubes, often due to anatomical factors or inflammation, can lead to fluid buildup and otitis media.

Allergies: Allergic reactions may contribute to inflammation and congestion in the Eustachian tubes, increasing the risk of otitis media.

Environmental Factors: Exposure to secondhand smoke or pollutants can be a risk factor for ear infections in children.

Otitis Externa (Swimmer's Ear):

Moisture and Humidity: Otitis externa is commonly associated with excess moisture in the ear canal, which creates an environment conducive to bacterial or fungal growth. This can be more prevalent in warm and humid conditions.

Trauma or Irritation: Scratching the ear canal with objects like cotton swabs or inserting foreign objects can damage the delicate skin of the ear canal, leading to inflammation.

Excessive Earwax: Buildup of earwax can create a favorable environment for bacterial growth and contribute to otitis externa.

Preventive measures for both types of otitis in children include practicing good hygiene, avoiding exposure to smoke, and addressing allergies when present. Prompt treatment of respiratory infections and avoiding prolonged exposure to water in the ear can help prevent ear infections.

If a child exhibits symptoms of ear infection, such as ear pain, drainage, or changes in hearing, it's important to consult a healthcare professional for proper diagnosis and treatment. Treatment may involve antibiotics for bacterial infections, pain management, and, in some cases, the placement of ear tubes for recurrent otitis media.

5. **How to interpret the test of tonal audiometry, what is the norm of hearing?**

Tonal audiometry is a hearing test that measures a person's ability to hear sounds at different frequencies and intensities. The results of tonal audiometry are typically presented on an audiogram, which is a graph that shows the individual's hearing thresholds at various frequencies. Here's how to interpret tonal audiometry results:

Audiogram Basics:

The horizontal axis of the audiogram represents frequencies, usually ranging from low to high frequencies (measured in Hertz, Hz).

The vertical axis represents intensity or loudness, measured in decibels (dB). The top of the audiogram represents loud sounds, and the bottom represents soft sounds.

Symbols on the Audiogram:

O (circle): Represents the right ear.

X (cross): Represents the left ear.

Audiometric Thresholds: The points on the graph where the symbols are plotted indicate the softest sounds that the individual can hear at each frequency. The lower the threshold, the better the hearing.

Degrees of Hearing Loss:

Normal Hearing: Thresholds at or below 20 dB across all frequencies are considered within the normal range.

Mild Hearing Loss: Thresholds between 20 and 40 dB.

Moderate Hearing Loss: Thresholds between 40 and 60 dB.

Severe Hearing Loss: Thresholds between 60 and 80 dB.

Profound Hearing Loss: Thresholds above 80 dB.

Configuration of Hearing Loss:

The shape or pattern of the audiogram can provide information about the configuration of hearing loss (e.g., flat, sloping, rising).

Speech Frequencies:

Important speech frequencies are typically tested at 500 Hz, 1000 Hz, 2000 Hz, 4000 Hz, and 8000 Hz.

It's important to note that the interpretation of tonal audiometry results should be done by a qualified audiologist or hearing healthcare professional. They will consider the entire audiogram, assess both ears, and provide appropriate recommendations based on the individual's hearing profile.

If you have a specific audiogram result and would like more detailed information, consulting with an audiologist is recommended, as they can provide personalized guidance based on the specific context and needs of the individual.

6. **What is the ABR test in a child?**

The Auditory Brainstem Response (ABR) test, also known as brainstem auditory evoked response (BAER), is a diagnostic test used to evaluate the auditory nerve and brainstem's responses to sound. It is commonly employed in both adults and children, including infants and newborns. In children, the ABR test is particularly useful when traditional behavioral hearing tests are challenging to conduct due to age or developmental factors.

Here's how the ABR test works:

Electrode Placement: Small electrodes are placed on the child's scalp and sometimes on the earlobes. These electrodes pick up electrical responses generated by the auditory nerve and brainstem in response to sound stimuli.

Stimuli Presentation: The child is exposed to a series of brief clicking sounds or tone bursts through earphones. These sounds are presented at different intensities and frequencies.

Measurement of Responses: The electrodes pick up the electrical responses generated by the auditory nerve and brainstem as a result of the presented sounds. The responses are then recorded and analyzed.

Threshold Determination: The ABR test can provide information about the child's hearing thresholds, which are the softest sounds they can detect.

The ABR test is valuable in several scenarios, including:

Newborn Hearing Screening: It is often used as part of newborn hearing screening programs to identify hearing issues early in infancy.

Young Children: When traditional behavioral testing is challenging due to a child's age or developmental stage.

Neurological Assessment: It can provide information about the integrity of the auditory nerve and brainstem pathways.

The ABR test is a non-invasive and objective measure of hearing function. While it does not provide information about the child's perception of speech or language development, it is an essential tool in the comprehensive evaluation of a child's auditory system. The results of the ABR test can help guide further diagnostic and intervention efforts if hearing issues are identified. A qualified audiologist typically administers and interprets the ABR test.

7. **What is newborn hearing screening?**

Newborn hearing screening is a set of tests designed to identify hearing issues in infants shortly after birth. The goal of newborn hearing screening is to identify hearing loss early so that intervention and treatment can begin promptly, promoting optimal language and speech development. Early detection is crucial because infants learn language and communication skills during their early months and years of life, and hearing is fundamental to this process.

There are two main types of newborn hearing screening tests:

Otoacoustic Emissions (OAE) Test:

In this test, a small probe is placed in the baby's ear, and it emits a sound. If the baby's inner ear (cochlea) is functioning properly, it will produce an echo-like response called an otoacoustic emission. The presence of these emissions suggests normal hearing.

Auditory Brainstem Response (ABR) Test:

This test measures the electrical activity of the auditory nerve and brainstem in response to sound. Small electrodes are placed on the baby's scalp, and the baby listens to clicking sounds. The test records the electrical responses, and the audiologist analyzes them to determine the baby's hearing status.

Newborn hearing screening is usually conducted in the hospital shortly after birth, often before the baby is discharged. The tests are quick, non-invasive, and typically do not cause discomfort to the infant. If the initial screening suggests a potential hearing issue, further diagnostic evaluations, such as a comprehensive audiological assessment, may be recommended.

It's important to note that not all babies who fail the initial screening have permanent hearing loss. Some infants may have temporary hearing issues due to factors like fluid in the ear or debris, and a follow-up evaluation helps differentiate between temporary and permanent hearing loss.

Early intervention and appropriate support for infants identified with hearing loss can significantly impact their language and communication development. If you have concerns about your baby's hearing, it's important to discuss them with healthcare providers, and if necessary, follow through with recommended hearing screening and evaluations.

8. **What is the course of exudative otitis media in a child?**

Exudative otitis media (EOM), also known as serous or secretory otitis media, is a condition characterized by the accumulation of fluid in the middle ear without signs of acute infection. This condition is commonly seen in children and can have various courses, depending on factors such as the underlying cause, treatment, and individual response. Here is a general overview of the course of exudative otitis media in a child:

Onset and Development:

EOM often develops as a result of dysfunction of the Eustachian tube, which connects the middle ear to the back of the throat. This dysfunction can lead to poor ventilation and drainage of the middle ear, causing fluid accumulation.

Asymptomatic or Mild Symptoms:

In some cases, children with EOM may not exhibit noticeable symptoms, or they may experience only mild symptoms such as a feeling of fullness in the ear, mild hearing loss, or occasional popping sounds.

Persistence of Fluid:

The fluid in the middle ear may persist for an extended period, leading to a chronic or recurrent condition. This persistence can be influenced by factors like allergies, exposure to secondhand smoke, or anatomical issues with the Eustachian tube.

Hearing Loss:

One of the primary concerns with EOM is its impact on hearing. The accumulation of fluid in the middle ear can interfere with the transmission of sound, resulting in conductive hearing loss.

Spontaneous Resolution:

In some cases, EOM may resolve spontaneously without intervention. As a child grows and the Eustachian tube matures, the ability to maintain proper ventilation and drainage of the middle ear may improve, leading to resolution of the condition.

Treatment Options:

When EOM persists or causes significant hearing loss, healthcare providers may consider interventions. Treatment options may include the management of underlying factors (such as allergies), placement of ear tubes (tympanostomy tubes) to allow fluid drainage, or other medical interventions.

Follow-up and Monitoring:

Children with EOM typically undergo regular follow-up evaluations to monitor the condition and assess hearing. Healthcare providers may adjust the treatment plan based on the child's progress.

It's important for parents and caregivers to work closely with healthcare providers to manage exudative otitis media effectively. Regular monitoring and appropriate interventions can help minimize the impact of EOM on a child's hearing and overall well-being.

9. **How can hearing be tested in a one-year-old child?**

Testing the hearing of a one-year-old child requires specialized techniques designed to accommodate their age and developmental stage. Here are two common methods used to assess hearing in infants and toddlers, including one-year-olds:

Behavioral Observation Audiometry (BOA):

This method involves observing the child's behavioral responses to sound. While the child is engaged in activities or play, the audiologist or healthcare professional presents sounds at different frequencies and volumes and monitors the child's reactions.

Responses may include the child turning their head toward the sound, displaying a startle reflex, or showing heightened attention. This method is suitable for children as young as a few months old.

Visual Reinforcement Audiometry (VRA):

VRA is a more structured method that involves pairing sound stimuli with a visual reward. The child is conditioned to associate a sound with a rewarding visual stimulus, such as a lighted toy or an animated display. When the child responds to the sound, they are rewarded visually.

This method is often used with children between six months and two years of age. The child is typically seated on a caregiver's lap, and speakers are used to present sounds from different directions.

It's important to note that testing the hearing of a one-year-old may require adaptation and flexibility based on the child's behavior and attention span. The testing environment is often designed to be child-friendly and engaging to encourage the child's participation.

If hearing concerns are identified during these initial screenings, more comprehensive testing, such as auditory brainstem response (ABR) testing, may be recommended. ABR is a neurophysiological test that measures the electrical activity of the auditory nerve and brainstem in response to sound.

Parents and caregivers play a crucial role in providing information about their child's responses to sound and should communicate any concerns to healthcare professionals. If there are suspected hearing issues or if the child is at risk for hearing loss, early intervention and further diagnostic testing may be recommended to address any potential hearing-related challenges.

10. **What is the verbal audiometry test?**

Verbal audiometry is a type of hearing test that assesses an individual's ability to understand and repeat spoken words or phrases. This test is commonly used to evaluate the functional aspects of hearing, particularly in terms of speech perception and discrimination. Verbal audiometry is often administered to individuals who are old enough to respond verbally, typically starting around age 5 or 6.

Here's an overview of how verbal audiometry is typically conducted:

Word Recognition Testing:

During a word recognition test, the individual listens to a list of single-syllable words or two-syllable words presented at a comfortable listening level. The words are usually spoken by the audiologist through headphones.

Scoring Responses:

The individual is then asked to repeat each word they hear. The audiologist records the individual's responses, noting which words were repeated correctly and which were misunderstood or not heard accurately.

Percentage of Correct Responses:

The results are expressed as a percentage of correct responses. This percentage reflects the individual's ability to accurately recognize and repeat the presented words.

Speech Discrimination:

In addition to word recognition testing, speech discrimination testing may be performed. This involves presenting sentences or phrases, and the individual is asked to repeat them. This type of testing provides information about the individual's ability to understand speech in a more complex context.

Verbal audiometry is an important component of a comprehensive audiological assessment, as it assesses how well an individual can understand and communicate with spoken language. The results of verbal audiometry, along with other tests like pure-tone audiometry and tympanometry, contribute to the overall understanding of a person's hearing abilities and may guide recommendations for intervention, such as hearing aids or other assistive devices.

It's important to note that verbal audiometry is typically used with individuals who have sufficient language and cognitive skills to participate in the test. For younger children or individuals with developmental or communication challenges, alternative methods, such as play audiometry or visual reinforcement audiometry, may be employed to assess hearing abilities.

11. **What types of tympanograms are distinguished in impedance audiometry?**

Tympanometry is a test used in impedance audiometry to assess the mobility of the eardrum (tympanic membrane) and the middle ear system. The test measures the compliance of the middle ear as air pressure in the ear canal is varied. Tympanograms are graphical representations of the results, and they help audiologists diagnose various middle ear conditions. There are several types of tympanograms, and they are categorized based on their shape and characteristics:

Type A Tympanogram:

This is considered a normal tympanogram. It shows a peak in compliance at the midpoint of the pressure range tested. The peak indicates that the eardrum and middle ear system are functioning well, and there is normal movement in response to changes in air pressure.

Type As Tympanogram:

A Type As tympanogram has a reduced peak compliance, suggesting reduced mobility of the tympanic membrane. This pattern is often associated with stiffening of the middle ear system, which can occur in conditions such as otosclerosis or partial ossicular discontinuity.

Type Ad Tympanogram:

A Type Ad tympanogram also has reduced peak compliance, but with an abnormally deep ear canal volume. This pattern is often associated with a flaccid or hypermobile tympanic membrane, which can be seen in conditions like a patulous Eustachian tube.

Type B Tympanogram:

A Type B tympanogram does not show a clear peak and is flat across the pressure range. This pattern can indicate a lack of mobility in the eardrum, often seen in conditions such as middle ear effusion (fluid in the middle ear) or a tympanic membrane perforation.

Type C Tympanogram:

A Type C tympanogram has a peak compliance shifted to the negative pressure side of the graph, indicating negative middle ear pressure. This pattern is often associated with Eustachian tube dysfunction or congestion, where the pressure in the middle ear is lower than normal.

These tympanogram types provide valuable information about the status of the middle ear, Eustachian tube function, and potential issues affecting hearing. Tympanometry is often performed as part of a comprehensive audiological evaluation to assess the overall health of the middle ear and contribute to the diagnosis and management of various ear conditions.

12. **For what purpose is an ear impression performed in children?**

Ear impressions, also known as earmolds or ear impressions, involve making a mold or impression of the ear's external shape and ear canal. These impressions are custom-made for an individual and are used for various purposes. In the context of children, ear impressions may be performed for the following purposes:

Hearing Aid Fittings:

Ear impressions are commonly used in the fitting of custom hearing aids for children. Custom hearing aids are designed to fit the unique shape and size of the child's ear, providing a comfortable and secure fit. The impressions allow the hearing healthcare professional to create hearing aids tailored to the child's specific ear anatomy.

Earmold for Hearing Protection:

Children may require hearing protection in certain noisy environments, such as concerts, sporting events, or loud recreational activities. Custom earmolds can be created based on ear impressions to provide a snug and effective fit, offering personalized hearing protection.

In-Ear Monitors for Musicians:

If a child is involved in music, particularly live performances, custom in-ear monitors can be created based on ear impressions. These monitors provide a comfortable and secure fit, delivering high-quality sound while also protecting the ears from excessive noise.

Swim Plugs or Water Protection:

Some children may benefit from custom swim plugs or water protection earmolds. These are designed to keep water out of the ears during activities such as swimming or bathing. The ear impressions help create a precise fit to prevent water from entering the ear canal.

Communication Earpieces:

In certain situations, such as for children with hearing loss who use communication devices, custom-molded earpieces may be needed. These earpieces can be created based on ear impressions to ensure a comfortable and effective fit for communication devices.

It's important to note that making ear impressions for children requires a gentle and child-friendly approach. Professionals, such as audiologists or hearing healthcare providers, with experience working with children, can ensure that the process is comfortable and stress-free for the child.

Overall, ear impressions for children serve a variety of purposes, all aimed at improving their auditory experiences, protecting their hearing, and enhancing their overall quality of life in various environments.

13. **How to determine air conduction in the audiogram?**

Air conduction testing in an audiogram is used to assess an individual's ability to hear sounds transmitted through the air. The audiogram is a graph that shows the softest sounds a person can hear at different frequencies (pitch) and intensities (volume). Air conduction testing involves the use of headphones or inserts, and the individual responds to various tones presented by the audiologist. Here's how air conduction testing is typically conducted:

Audiometric Equipment:

The audiologist uses specialized audiometric equipment, including calibrated headphones or inserts and an audiometer. The audiometer generates pure tones at different frequencies and intensities.

Frequency Selection:

The audiologist selects specific frequencies to be tested. Standard audiometric frequencies include 250 Hz, 500 Hz, 1000 Hz, 2000 Hz, 4000 Hz, and 8000 Hz. These frequencies are measured in Hertz (Hz).

Intensity Levels:

For each selected frequency, the audiologist presents tones at different intensity levels, measured in decibels hearing level (dB HL). The intensity starts at a level that is typically easy to hear, and then decreases in 5 or 10 dB steps until the softest sound that the individual can hear is determined.

Patient Response:

The individual being tested responds to each tone by pressing a button, raising a hand, or providing another predetermined signal when they hear the sound. The responses are recorded by the audiologist.

Plotting the Audiogram:

The results are plotted on an audiogram, with frequency on the horizontal axis and intensity (dB HL) on the vertical axis. The symbols on the audiogram represent the softest sounds the individual can hear at each tested frequency.

Threshold Determination:

The softest intensity level at which the individual can hear each frequency is known as the hearing threshold. The hearing thresholds are used to assess the degree of hearing loss, if any, at different frequencies.

Air conduction testing assesses the entire auditory system, from the outer ear to the inner ear and beyond. It provides information about the individual's overall hearing sensitivity and helps identify the type and configuration of hearing loss. If a hearing loss is identified, further testing, such as bone conduction testing, may be conducted to determine the specific site of the hearing impairment within the auditory system.

14. **What should be done in the case of an abnormal result of hearing screening in a newborn?**

If a newborn's hearing screening results are abnormal, it is important to follow up with additional testing and evaluation promptly. Early detection of hearing loss is crucial for intervention and support to ensure optimal language and communication development. Here are the general steps to be taken when an abnormal result is obtained in a newborn hearing screening:

Repeat Screening:

In some cases, a false positive result may occur due to factors such as debris in the ear canal or the infant being unsettled during the initial screening. Before proceeding with additional testing, it is common to repeat the hearing screening to confirm the result.

Diagnostic Auditory Brainstem Response (ABR) Testing:

If the repeat screening still indicates a potential hearing issue, the next step is often a diagnostic Auditory Brainstem Response (ABR) test. ABR is a more comprehensive and specific test that measures the electrical activity of the auditory nerve and brainstem in response to sound. It provides information about the presence, degree, and type of hearing loss.

Medical Evaluation:

If the ABR test confirms hearing loss, the infant should undergo a medical evaluation by an otolaryngologist (ear, nose, and throat specialist) or a pediatric audiologist. The medical evaluation helps identify any underlying medical conditions or anatomical issues that may be contributing to the hearing loss.

Intervention Planning:

Once the type and degree of hearing loss are determined, an intervention plan can be developed. This plan may include recommendations for hearing aids, cochlear implants (if appropriate), and early intervention services to support the child's communication and language development.

Family Support and Education:

Supporting the family is an essential aspect of managing hearing loss in a newborn. Families should receive information and resources to help them understand the implications of hearing loss and how to support their child's communication and development. Educational programs and resources for families with children who are deaf or hard of hearing are often available.

Follow-up Monitoring:

Regular follow-up appointments and monitoring are crucial to assess the child's progress and make any necessary adjustments to the intervention plan. Ongoing audiological evaluations help track changes in hearing and ensure that appropriate interventions are in place as the child grows.

It's important for parents and caregivers to actively participate in the process, communicate openly with healthcare professionals, and seek support from relevant services and organizations specializing in early intervention for hearing loss. Early identification and intervention significantly improve outcomes for children with hearing loss.

15. **What do the yellow and blue certificates in the child's health book mean in the context of hearing screening of children in Poland?**

As of my last knowledge update in January 2022, I do not have specific information about the color-coding system used in child health books in Poland for hearing screening certificates. Health systems and administrative procedures can evolve, and it's possible that there have been changes or updates since then.

In many countries, including Poland, health books or child health records often contain information related to the child's health, vaccinations, growth, and development milestones. Hearing screening is an essential component of early childhood health monitoring, and certificates may be issued to document the results of hearing screening tests.

To obtain accurate and up-to-date information about the specific color-coding system used for hearing screening certificates in Poland, I recommend checking with local health authorities, pediatricians, or the relevant government health agencies. They can provide details about the current practices, protocols, and the meaning of different colors or codes on hearing screening certificates in the context of child health records in Poland.

16. **What are the contraindications to the use of air-conduction hearing aids?**

Air-conduction hearing aids are commonly used to amplify sounds and improve hearing for individuals with hearing loss. While they are generally safe and effective, there are specific situations or conditions where the use of air-conduction hearing aids may not be advisable. Contraindications to the use of air-conduction hearing aids include:

Presence of Acute Ear Infections:

Individuals with active or acute ear infections, such as otitis media with drainage, may be advised to avoid using hearing aids until the infection is treated and resolved. Introducing hearing aids in the presence of an active infection can potentially worsen the condition.

Severe External Otitis or Dermatitis:

Severe external otitis (inflammation of the ear canal) or dermatitis of the ear may be a contraindication to using traditional air-conduction hearing aids. The presence of inflammation or open sores may make wearing hearing aids uncomfortable or exacerbate the existing condition.

Drainage or Otitis Externa with Inflammation:

Conditions involving persistent drainage or inflammation of the ear canal, such as chronic otitis externa, may be contraindications to using air-conduction hearing aids. These conditions may interfere with the proper functioning and fit of hearing aids.

Anatomical Abnormalities or Deformities:

Some individuals may have anatomical abnormalities or deformities of the ear that make it challenging to achieve a proper fit with conventional hearing aids. In such cases, alternative hearing solutions may be explored.

Allergic Reactions:

Allergic reactions to materials used in hearing aids (e.g., the casing, ear molds, or adhesives) may be a contraindication. In these cases, individuals may need hypoallergenic materials or different hearing aid options.

Inability to Tolerate Hearing Aid Use:

Some individuals may experience discomfort, irritation, or a sense of being overwhelmed when using hearing aids. In such cases, it's important to address the specific issues causing discomfort and explore alternative solutions or adjustments.

It's crucial for individuals with hearing loss to undergo a comprehensive hearing evaluation by an audiologist or hearing healthcare professional. The audiologist can assess the individual's specific needs, determine the type and degree of hearing loss, and recommend suitable hearing solutions based on the individual's overall health and anatomical considerations.

Ultimately, the decision to use air-conduction hearing aids or explore alternative solutions depends on the individual's unique circumstances and the guidance of qualified healthcare professionals.

17. **What are the most effective methods of treatment of exudative otitis media in a child?**

Exudative otitis media (EOM), also known as serous or secretory otitis media, is a condition characterized by the accumulation of fluid in the middle ear without signs of acute infection. The management of EOM in children often depends on the underlying cause and the presence of associated factors. Here are some methods of treatment that may be considered for exudative otitis media in a child:

Observation and Monitoring:

In some cases, especially if the fluid accumulation is mild and does not significantly impact the child's hearing or quality of life, a healthcare professional may recommend a period of observation and monitoring. EOM can resolve on its own over time.

Management of Underlying Factors:

Addressing underlying factors contributing to EOM can be important. For example, managing allergies or respiratory infections may help reduce inflammation and fluid buildup in the middle ear.

Nasal Steroids:

Nasal steroids may be recommended to reduce nasal congestion and inflammation, especially if EOM is associated with conditions like allergic rhinitis. These medications can help improve Eustachian tube function and ventilation of the middle ear.

Antibiotics:

While EOM is not typically caused by bacterial infection, if there is evidence of bacterial infection or if EOM is associated with acute otitis media, a healthcare professional may prescribe antibiotics. Antibiotics are not the primary treatment for EOM, but they may be used in specific cases.

Autoinflation (Valsalva Maneuver):

In some cases, healthcare providers may recommend autoinflation exercises, also known as the Valsalva maneuver. This involves encouraging the child to perform actions that help equalize pressure in the middle ear, such as blowing up a balloon.

Tympanostomy Tubes (Ear Tubes):

For recurrent or persistent cases of EOM, where fluid continues to accumulate and impact hearing, healthcare providers may recommend the insertion of tympanostomy tubes. These tiny tubes are placed through the eardrum to allow ventilation and drainage of the middle ear.

Myringotomy:

Myringotomy is a surgical procedure where a small incision is made in the eardrum to drain fluid from the middle ear. This may be performed in conjunction with the placement of tympanostomy tubes.

Treatment decisions are individualized based on the child's specific circumstances, the severity of symptoms, and the duration of the condition. It's important for parents or caregivers to work closely with healthcare professionals to determine the most appropriate treatment plan for their child's exudative otitis media. Regular follow-up visits are typically recommended to monitor the child's progress and adjust the treatment plan as needed.

18. **What is the result of a verbal audiometry test and how to interpret it?**

Verbal audiometry is a type of hearing test that assesses an individual's ability to understand and repeat spoken words or phrases. The results of a verbal audiometry test are typically presented in the form of a percentage score, indicating the percentage of words or phrases correctly repeated by the individual being tested.

Here's how to interpret the results of a verbal audiometry test:

Word Recognition Score (WRS):

The percentage score obtained in a verbal audiometry test is often referred to as the Word Recognition Score (WRS) or Speech Discrimination Score. It represents the proportion of words or phrases presented to the individual that were correctly identified and repeated.

Percentage Ranges:

The percentage can range from 0% to 100%, with 100% indicating perfect word recognition. The results are typically categorized into different ranges:

Normal Range (90-100%): Excellent word recognition.

Mild Hearing Loss (80-89%): Slight difficulty with word recognition.

Moderate Hearing Loss (70-79%): Moderate difficulty with word recognition.

Severe Hearing Loss (50-69%): Significant difficulty with word recognition.

Profound Hearing Loss (<50%): Poor word recognition.

Configuration of Hearing Loss:

The pattern of word recognition scores can provide information about the configuration of hearing loss. For example, if there is a significant difference in scores across different frequencies, it may suggest a specific pattern of hearing loss.

Use of Phonetically Balanced Word Lists:

Verbal audiometry tests often use phonetically balanced word lists, which include words that contain a representative sample of speech sounds. This helps ensure that the test accurately reflects an individual's ability to understand everyday speech.

Consideration of Context:

It's important to consider the overall context of the individual's hearing ability. For example, someone with a mild hearing loss may still have good word recognition in a quiet environment but may struggle more in noisy or challenging listening situations.

Interpreting the results of a verbal audiometry test should be done by a qualified audiologist or hearing healthcare professional. They will consider the entire audiometric profile, including pure-tone audiometry results and other relevant tests, to provide a comprehensive understanding of the individual's hearing abilities. The results help guide decisions regarding the need for hearing aids, assistive devices, or other interventions to address communication difficulties associated with hearing loss.

19. **What are the results of otoscopy for otosclerosis?**

Otoscopy alone may not provide definitive results for diagnosing otosclerosis, as it is primarily a clinical examination technique to visualize the external ear canal and the eardrum. Otosclerosis is an inner ear disorder that involves abnormal bone growth in the middle ear, particularly around the stapes bone, which can lead to hearing loss.

To diagnose otosclerosis, a combination of clinical assessments and diagnostic tests is typically used. Here are some aspects of otoscopy and related tests that may be relevant in the context of otosclerosis:

Otoscopy:

During otoscopy, the healthcare provider examines the ear canal and eardrum using an otoscope. Signs that may be observed in otoscopy include a normal or slightly retracted eardrum, and sometimes a bluish hue (Schwartz sign) may be visible due to vascular changes associated with otosclerosis. However, otoscopy alone cannot definitively diagnose otosclerosis.

Audiometry:

Pure-tone audiometry is a key diagnostic test for otosclerosis. Individuals with otosclerosis often exhibit a characteristic pattern of conductive hearing loss, particularly at low frequencies. The air-bone gap on the audiogram may be indicative of otosclerosis, where bone conduction thresholds are within the normal range, but air conduction thresholds are elevated.

Tympanometry:

Tympanometry measures the mobility of the eardrum and middle ear system. In otosclerosis, tympanometry may show a characteristic pattern, such as a normal compliance but a reduced ear canal volume. Tympanometry, along with audiometry, contributes to the diagnosis of otosclerosis.

Stapedial Reflex Testing:

Stapedial reflex testing involves measuring the reflex contraction of the stapedius muscle in response to loud sounds. Individuals with otosclerosis may have absent or elevated stapedial reflex thresholds.

Imaging Studies:

High-resolution imaging techniques like temporal bone CT scans can provide detailed images of the middle ear structures. Imaging may reveal specific features associated with otosclerosis, such as the characteristic "double-ring" appearance around the stapes on a CT scan.

Ultimately, the diagnosis of otosclerosis is often made based on a combination of clinical history, physical examination, audiometric testing, and imaging studies. If otosclerosis is suspected, an otolaryngologist or an ear, nose, and throat (ENT) specialist may be consulted for further evaluation and management. Surgical intervention, such as stapedectomy, is a common treatment option for individuals with symptomatic otosclerosis.

20. **What implantable hearing prostheses can be used in children?**

Several implantable hearing prostheses can be used in children with hearing loss. The choice of a specific device depends on factors such as the type and degree of hearing loss, the child's age, and individual factors. Here are some of the implantable hearing prostheses commonly used in children:

Cochlear Implants:

Cochlear implants are electronic devices that are surgically implanted to provide direct electrical stimulation to the auditory nerve. They are suitable for children with severe to profound sensorineural hearing loss who do not benefit significantly from traditional hearing aids. Cochlear implants can be used in infants and older children.

Bone-Anchored Hearing Aids (BAHA):

Bone-Anchored Hearing Aids (BAHA) are surgically implanted devices that use bone conduction to transmit sound vibrations directly to the inner ear. They are suitable for children with conductive or mixed hearing loss or single-sided deafness. BAHA devices can be implanted in older children once the skull has matured, and there are also options for non-surgical BAHA solutions.

Auditory Brainstem Implants (ABI):

Auditory Brainstem Implants (ABI) are implanted devices designed for individuals with a missing or damaged cochlear nerve, which may occur in cases of bilateral cochlear nerve deficiency. ABIs are more commonly used in children with neurofibromatosis type II (NF2).

Middle Ear Implants:

Middle ear implants are surgically placed in the middle ear and are designed to enhance sound transmission through the natural hearing pathway. These devices can be considered for certain types of sensorineural or mixed hearing loss in children.

Vibrant Soundbridge (VSB):

The Vibrant Soundbridge is a type of middle ear implant that directly drives the middle ear structures. It is suitable for children with sensorineural or mixed hearing loss and can be implanted in children as young as 5 years old.

Earlens Contact Hearing Solution:

The Earlens Contact Hearing Solution is a non-surgical, light-driven hearing aid system that uses a lens placed on the eardrum to directly vibrate it. This system is suitable for children with mild to moderately severe sensorineural hearing loss.

The selection of an implantable hearing prosthesis is based on a thorough evaluation by an audiologist, otolaryngologist (ENT specialist), and other relevant healthcare professionals. The child's age, hearing profile, medical history, and individual needs are carefully considered to determine the most appropriate device.

It's important for parents and caregivers to work closely with the healthcare team to make informed decisions about the type of implantable hearing prosthesis that will best meet the child's needs and provide optimal auditory outcomes.
